# Depression is Associated with Treatment Response Trajectories in Adults with Prolonged Grief Disorder: A Machine Learning Analysis

**DOI:** 10.1101/2024.12.11.24318861

**Authors:** Adam Calderon, Matthew Irwin, Naomi M. Simon, M. Katherine Shear, Christine Mauro, Sidney Zisook, Charles F. Reynolds, Matteo Malgaroli

## Abstract

Although evidence-based treatments for Prolonged Grief Disorder (PGD) exist, pretreatment characteristics associated with differential improvement trajectories have not been identified. To identify clinical factors relevant to optimizing PGD treatment outcomes, we used unsupervised and supervised machine learning to study treatment effects from a double-blinded, placebo-controlled, randomized clinical trial. Participants were randomized into four treatment groups for 20 weeks: citalopram with grief-informed clinical management, citalopram with prolonged grief disorder therapy (PGDT), pill placebo with PGDT, or pill placebo with clinical management. The trial included 333 PGD patients aged 18–95 years (*M_age_* = 53.9; *SD* ± 14.4), predominantly female (77.4%) and white (84.4%). Symptom trajectories were assessed using latent growth mixture modeling based on Inventory for Complicated Grief scores collected at six time points every 4 weeks. The relationship between patient-level characteristics and assigned trajectories was examined using logistic regression with elastic net regularization based on the administration of citalopram, PGDT, and risk factors for developing PGD. Three distinct response trajectories were identified: lesser severity responders (60%, n = 200), greater severity responders (18.02%, n = 60), and non-responders (21.92%, n = 73). Differences between greater severity responders and non-responders emerged as statistically significant by Week 8. The elastic net model demonstrated acceptable discrimination between responders and non-responders (AUC = .702; accuracy = .684). Higher baseline depression severity, grief-related functional impairment, and absence of PGDT were associated with reduced treatment response likelihood. These findings underscore the importance of early identification of clinical factors to optimize individualized PGD treatment strategies.

**Trial Registration:** clinicaltrials.gov Identifier: NCT01179568.

## 1. Introduction

With the inclusion of prolonged grief disorder (PGD) (Simon and Shear, 2024) as a new diagnosis in DSM-5-TR and ICD-11, there is a need to establish guidelines for matching patients to interventions to optimize treatment response. Among interventions studied for PGD to date, PGD therapy (PGDT) has been demonstrated to be particularly effective (Shear et al., 2016, 2014, 2005). In contrast, in the first double-blinded, placebo-controlled randomized trial (RCT) evaluating the efficacy of citalopram alone or in conjunction with PGDT for PGD, Shear et al. (2016) found no difference in response rates between citalopram and placebo. PGDT, compared to no PGDT, however, exhibited markedly better response. Moreover, combining citalopram with PGDT reduced co-occurring depressive symptoms but not PGD symptoms compared to PGDT plus placebo. Given the substantial impact of PGD on distress, impairment (Bonanno and Malgaroli, 2020), and suicidal ideation (Simon and Shear, 2024), it is imperative to identify patients not likely to respond to interventions – especially medication or PGD-informed support (Szuhany et al., 2021).

Machine learning methods offer the capacity to cluster individuals based on symptom patterns while suspending expectations of predetermined outcomes. This approach offers novel ways to utilize patient characteristics to examine propensity for different treatment outcomes (Malgaroli et al., 2022). To our knowledge, only one prior study has utilized such an approach to discriminate between resilience and symptomatic trajectories among bereaved adults (Schultebraucks et al., 2021). While informative, this study used depression symptom trajectories and did not assess PGD symptoms. We sought to explore whether machine learning methods may 1) identify clinically meaningful subgroups of grief improvement patterns, 2) determine at what point during the course of treatment response is distinguished from non-response, and 3) assess patient characteristics associated with differential trajectories of improvement in the first placebo-controlled trial of antidepressant pharmacotherapy, with and without PGDT, among patients with PGD (Shear et al., 2016).

## 2. Methods

### Participants and Setting

The current study analyzes clinical records from HEAL (Healing Emotions After Loss), a 20-week, four-site, double-blinded, placebo-controlled RCT of citalopram with and without PGDT for adults (18-95) with PGD conducted between March 2010 and September 2014 (Shear et al., 2016). All procedures were approved by the institutional review boards from each site, and all participants signed written informed consent before baseline assessment. Participants (N = 395) were bereaved individuals with clinician-confirmed primary PGD based on the Structured Clinical Interview for Complicated Grief (SCI-CG) (Bui et al., 2015a). Those with a current substance use disorder (past six months), a lifetime history of psychotic disorder, bipolar I disorder, active suicidal plans requiring hospitalization, a Montreal Cognitive Assessment score of less than 21, or a pending lawsuit or disability claim related to the death, as well as those undergoing concurrent psychotherapy or treatment with an antidepressant were excluded.

Participants were randomized to either citalopram with grief-informed clinical management (n = 101), citalopram with PGDT (n = 99), pill placebo plus PGDT (n = 96), or pill placebo with PGD-informed clinical management (n = 99) and were stratified by site and presence or absence of current major depressive disorder (MDD). PGDT was delivered in a manualized, 16-session format to facilitate adaptation in alignment with the dual process model of coping with bereavement by focusing on loss and restoration (Stroebe and Schut, 2010). Pharmacotherapists administering citalopram or placebo also provided PGD-informed clinical management.

### Measures

The outcome for our study was grief symptom severity as measured by the Inventory for Complicated Grief (ICG) (Prigerson et al., 1995). The ICG is a 19-item adult self-report questionnaire with responses rated on a 5-point frequency scale with total scores ranging from 0-76, with higher scores denoting greater severity. PGD symptoms were measured with the ICG at the first treatment visit and then 4, 8, 12, 16, and 20 weeks after. An ICG score of 30 was used as a clinical threshold of PGD for HEAL, in line with other studies (Shear et al., 2005; Zisook et al., 2018). The ICG holds excellent internal consistency (Cronbach’s *α* = .94), test-retest reliability (0.80), and good convergent validity with other grief symptom measures (Bui et al., 2015b; Prigerson et al., 1995). Additional clinical symptoms collected at baseline include depression (Quick Inventory Depressive Symptoms; QIDS-SR) (Rush et al., 2003), sleep disturbance (item 6 of the Pittsburgh Sleep Quality Index; PSQI) (Buysse et al., 1989), grief-related avoidance (Grief-Related Avoidance Questionnaire; GRAQ) (Shear et al., 2007), grief-related functional impairment (using the Work and Social Adjustment scale; WSAS) (Mundt et al., 2002), as well as social support (Interpersonal Support Evaluation List-Short Form; ISEL) (Merz et al., 2014).

### Data Analysis

Treatment response was analyzed with a three-step approach using data from all four treatment arms. Statistical analyses were performed in Mplus Version 7.4 and R Version 4.3.1 via the nestedcv package (Lewis et al., 2023; Muthén and Muthén, 2012; R Core Team, 2022). The current study followed the Transparent Reporting of a multivariable prediction model for Individual Prognosis Or Diagnosis (TRIPOD) + AI guidelines (Collins et al., 2024). Extended methods are provided in the **online supplement**.

#### Trajectories

First, we identified symptom improvement trajectories using unsupervised machine learning (latent growth mixture modeling; LGMM) (Muthén and Muthén, 2012). For LGMM development, ICG data from baseline to weeks 4, 8, 12, 16, and 20 after the first treatment visit were used. To lower the risk of modeling spurious subpopulations, we chose to run analyses only with those who reported two or more ICG assessments (n = 333). An intent-to-treat approach using Full Information Maximum Likelihood was used to handle missing ICG scores during LGMM estimation. LGMM does not rely on the assumption that a homogeneous mean response can meaningfully describe individuals; instead, LGMM allows to tease out subpopulations (or classes) characterized by discrete growth patterns (i.e., heterogeneous reactions to the loss). After the best fitting solution was found, a one-sample *t*-test analyzed differences between trajectories in ICG scores at each time point.

#### Treatment outcomes model

Second, we analyzed trajectory outcomes using supervised machine learning (logistic regression with elastic net regularization) (Zou and Hastie, 2005a). Elastic net regularization (elastic net) is a supervised machine learning method and a form of conventional regression that combines both ridge and lasso norms to provide a penalization term to balance stability and parsimony. We employed elastic net, given its ability to constrain coefficients among collinear variables and minimize model overfitting, with the lambda hyperparameter determining the magnitude and the alpha hyperparameter regulating the balance between the two norms (Zou and Hastie, 2005b). Assigned trajectories were dichotomized into two binary groups by combining the lesser severity and greater severity responders to identify clinical factors that distinguished non-responders. As candidate factors, we used demographic variables (age, gender, ethnicity, race, level of education), citalopram and PGDT administration, comorbid psychiatric diagnoses (major depressive disorder, generalized anxiety disorder, posttraumatic stress disorder with presence or absence of suicidal ideation), and baseline clinical symptoms (see measures) and loss characteristics (relation to bereaved [spouse, child], cause of death [sudden, violent], and time since loss [years since death]) considered risk factors for developing PGD (Szuhany et al., 2021). To optimize the probability of unbiased results, we utilized 5 x 5-fold nested cross-validation to prevent over-fitting (Lewis et al., 2023). Random forest imputation was performed in training and testing outer folds independently without knowledge of the outcome variable to avoid data leakage of information about the variable distribution between folds within the nested cross-validation. Synthetic minority oversampling technique (Chawla et al., 2002) was applied only on the outer training folds, given that balancing the whole dataset prior to outer cross-validation leads to data leakage of outer cross-validation hold-out samples into the outer training folds (Lewis et al., 2023). For model evaluation, we used Receiver Operating Characteristic (ROC) curves and Area Under the Curve (AUC) metrics to assess classification performance, with AUC values indicating discrimination quality (Hosmer et al., 2013).

#### Treatment Improvement Predictors

Third, we assessed the relative importance of each feature in the model based on their likelihood of increasing treatment response as determined by methods for explainable machine learning (SHapley Additive exPlanations; SHAP) (Lundberg and Lee, 2017). In sum, SHAP is based on the concept of Shapley values from cooperative game theory (Shapley and Snow, 1952). SHAP values assign a value to each feature that represents the average contribution of that feature across all possible combinations of features. The average SHAP value across all individuals is 0, but the average absolute SHAP value informs about relative feature importance. Thus, by considering all possible combinations, SHAP values provide a more comprehensive understanding of each feature’s contribution to the model’s predictions.

## 3. Results

Baseline characteristics are presented in **Table 1**. LGMM identified 3 treatment trajectories (n = 333; **Figure 1**): lower baseline ICG severity with marked improvement (60%, n = 200), greater baseline ICG severity with marked improvement (18.02%, n = 60), and greater baseline ICG severity non-responders (21.92%, n = 73). Differences in treatment response between the two groups with greater baseline severity were statistically significant by Week 8 (**Table 2**; *X^2^*(6, 333) = 26.867, *p* < .001). The elastic net model indicated an acceptable classification performance discriminating treatment response versus non-response trajectories with an AUC of .702 (95% CI, .597 - .722) (n = 333; accuracy, .684, precision, .867; recall, .703; specificity, .616; F1, .777). The 7 most important features identified using SHAP are shown in **Figure 2**. Characteristics associated with decreased probability of response were higher baseline depression severity (mean |SHAP| = 0.269), grief-related functional impairment (mean |SHAP| = 0.266), suicidal ideation (mean |SHAP| = 0.103), grief-related avoidance (mean |SHAP| = 0.056), and sleep disturbances (mean |SHAP| = 0.045). Receiving PGDT (mean |SHAP| = 0.209) and longer time since loss (mean |SHAP| = 0.081) were associated with increased probability of response. Sensitivity analyses separately examining response likelihood for PGDT and citalopram treatment arms are in the **online supplement**.

**Table 1.** Baseline socio-demographic and clinical characteristics of study sample

| **Features** |  | **Total**<br>**(n = 333)** |
| --- | --- | --- |
| *Categorical features* |  | **No. (%)** |
| Gender | Male | 75 (22.5) |
|  | Female | 257 (77.2) |
| Ethnicity | Non-Hispanic | 294 (88.3) |
|  | Hispanic or Latino | 38 (11.4) |
| Race | American Indian or Alaska Native | 6 (1.8) |
|  | Asian | 6 (1.8) |
|  | Black or African American | 31 (9.3) |
|  | Native Hawaiian or Other Pacific Islander | 1 (0.3) |
|  | White | 278 (83.5) |
|  | Did not wish to provide | 11 (3.3) |
| Education | Grade 7-12 | 3 (0.9) |
|  | High School/GED | 33 (9.9) |
|  | Part college | 90 (27.0) |
|  | Graduated 2-year college | 27 (8.1) |
|  | Graduated 4-year college | 69 (20.7) |
|  | Part graduate/professional school | 30 (9.0) |
|  | Completed graduate/professional school | 81 (24.3) |
| Marital status | Never married | 77 (23.1) |
|  | First marriage | 51 (15.3) |
|  | Remarried after divorce | 23 (6.9) |
|  | Remarried after widowed | 5 (1.5) |
|  | Separated | 5 (1.5) |
|  | Divorced, not currently married | 48 (14.4) |
|  | Widowed, not currently married | 124 (37.2) |
| Person who died | Husband | 86 (25.8) |
|  | Wife | 29 (8.7) |
|  | Significant Other | 13 (3.9) |
|  | Father | 29 (8.7) |
|  | Mother | 64 (19.2) |
|  | Son | 49 (14.7) |
|  | Daughter | 17 (5.1) |
|  | Brother | 16 (4.8) |
|  | Sister | 10 (3.0) |
|  | Grandfather | 1 (0.3) |
|  | Grandmother | 4 (1.2) |
|  | Other relative | 3 (0.9) |
|  | Friend | 10 (3.0) |
|  | Other | 1 (0.3) |
| Cause of death | Illness < 1 month | 66 (19.8) |
| | Illness ≥ 1 month | 152 (45.6) |
|  | Accident | 47 (14.1) |
|  | Murder | 16 (4.8) |
|  | Suicide | 44 (13.2) |
|  | Other | 8 (2.4) |
| Current PTSD<sup>a</sup> | Yes | 125 (37.5) |
| Current GAD<sup>a</sup> | Yes | 146 (43.8) |
| Current MDD<sup>a</sup> | Yes | 223 (67.0) |
| Suicidal ideation<sup>a</sup> | Yes | 96 (28.8) |
| Sleep disturbance [PSQI item] | Yes | 151 (45.3) |
| ***Continuous features*** |  | **mean (SD)** |
| Age |  | 53.9 (14.4) |
| Time since loss [years] |  | 4.8 (7.6) |
| Social support [ISEL] |  | 10.2 (3.2) |
| Grief-related avoidance [GRAQ] |  | 22.1 (12.9) |
| Depression symptoms [QIDS-SR] |  | 13.4 (4.0) |
| Grief-related functional impairment [WSAS] |  | 22.0 (9.7) |

*Note.* <sup>a</sup> = clinician-rated assessment at baseline. Abbreviations: PTSD, post-traumatic stress disorder; GAD, generalized anxiety disorder; MDD, major depressive disorder; PSQI item, item 6 of Pittsburgh Sleep Quality Index; ISEL, Interpersonal Support Evaluation List-Short Form; GRAQ, Grief-related Avoidance Questionnaire; QIDS-SR, The Quick Inventory of Depressive Symptomatology Self-Report; WSAS, Work and Social Adjustment scale.

**Figure 1.**
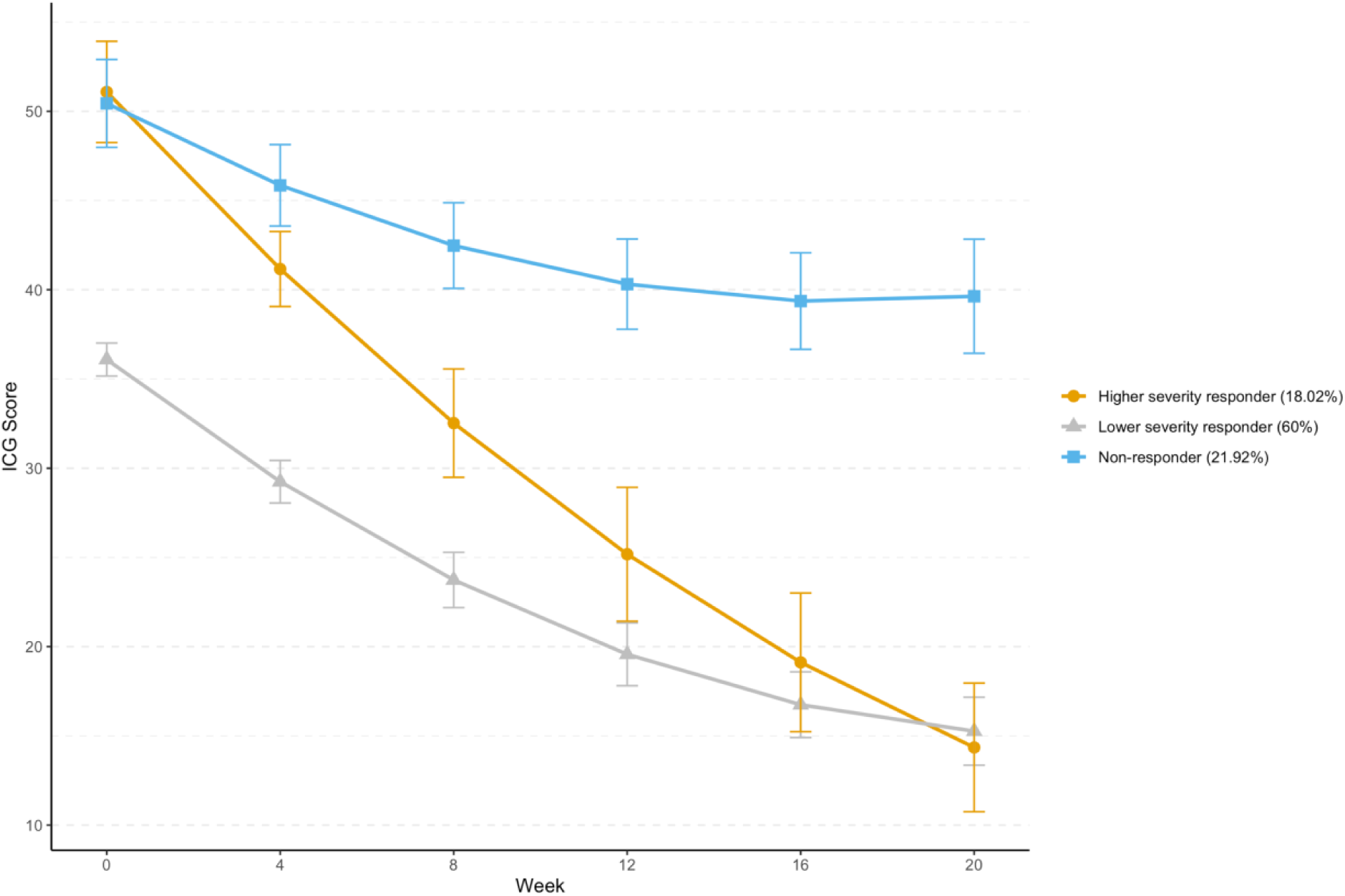
Treatment trajectories based on PGD severity. *Note*. n = 333. Each point represents the mean and 95% confidence interval of the Inventory of Complicated Grief (ICG) score for each trajectory at four-weeks intervals throughout treatment. An ICG score of 30 or greater is considered a positive screen for Prolonged Grief Disorder.

**Table 2.** Distribution of three treatment trajectory groups across four treatment arms

|  |  | Trajectory Group |  |  | Total |
| --- | --- | --- | --- | --- | --- |
|  |  | Lesser severity<br>responders | Greater severity<br>responders | Non-responders |  |
| **Tx**<br>**arm** | CIT + PGDT | 49 | 22 | 15 | 86 |
|  | PLA + PGDT | 49 | 26 | 13 | 88 |
|  | CIT + no<br>PGDT | 56 | 4 | 22 | 82 |
|  | PGDT + no<br>PGDT | 46 | 8 | 23 | 77 |
|  | Total | 200 | 60 | 73 | 333 |

*Note.* n = 333. Abbreviations: CIT, received citalopram; PGDT, Prolonged Grief Disorder therapy; PLA, pill placebo. A chi-square test indicated a significant relationship between treatment arms and trajectory classes, $\chi^2(6, 333) = 26.867, p = < .001$ .

**Figure 2.**
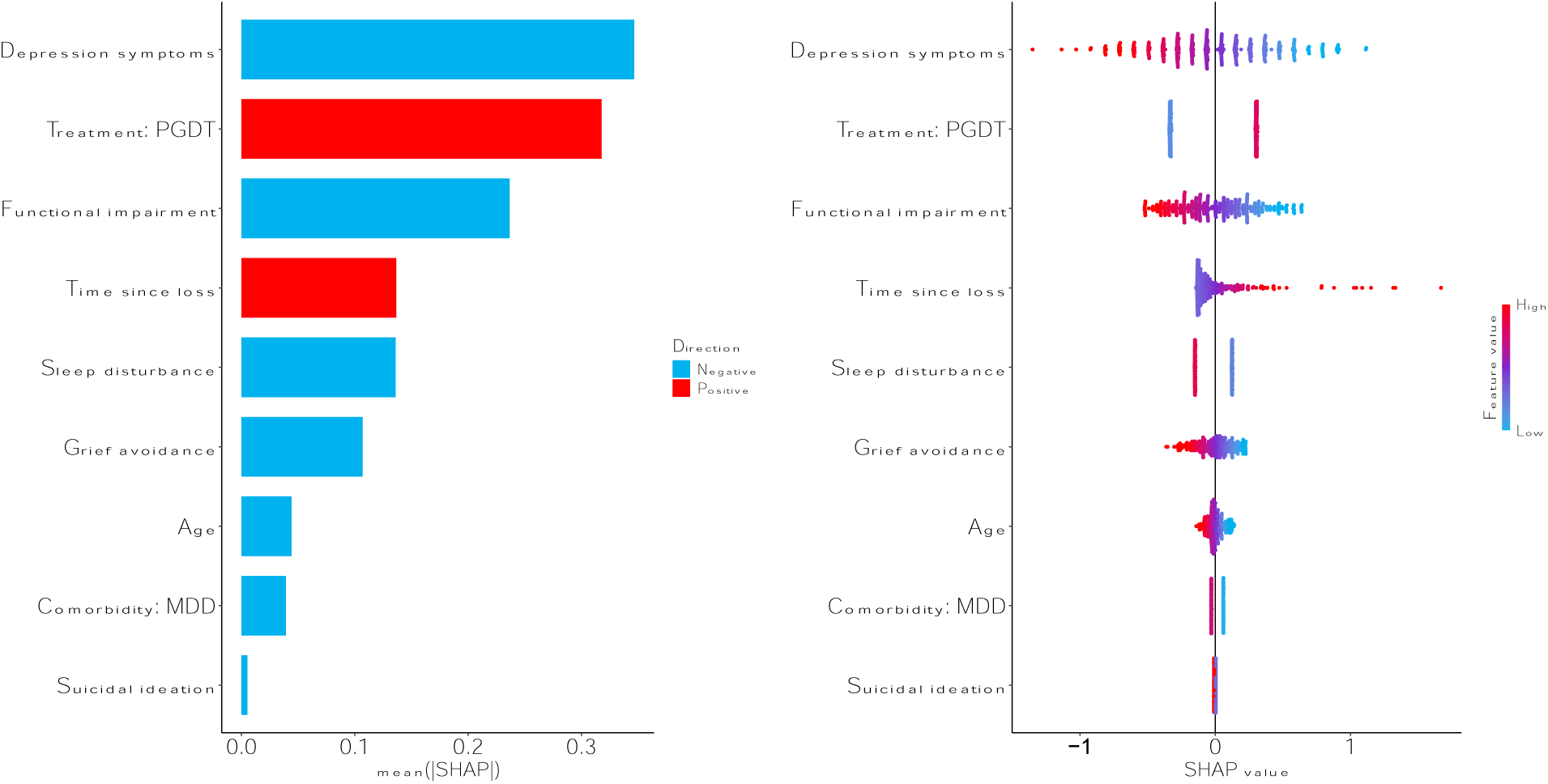
Variable importance and directionality based on SHapley Additive exPlanations (SHAP) *Note.* n = 333. The larger the SHAP value, the more important the model feature is in discriminating treatment response. The bar plot (left) shows the mean absolute SHAP value per feature. The bee swarm plot (right) displays every individual in the sample as a dot, to further illustrate the attribution value for each feature from low (blue) to high (red). Features are sorted by their global impact (*y* axis) and are arranged in descending order based on the magnitude of their total contribution. The (*x*-axis) represents the contribution to treatment response as log odds.

## 4. Discussion

We analyzed an RCT of citalopram and PGDT to answer three questions: (1) Can differential improvement trajectories in adults with PGD be identified? (2) At what point in treatment can response be distinguished from non-response? (3) Across treatment groups, are baseline patient characteristics associated with differential PGD treatment outcomes? Our results found three trajectories of ICG symptom severity: (a) moderate level baseline ICG severity showing improvement in ICG score to a putative nonclinical level; (b) high baseline ICG severity showing marked improvement in ICG score to a putative nonclinical level; and (c) high baseline ICG severity without clinical improvement throughout treatment. The two elevated baseline symptom trajectories diverged significantly into response and non-response by Week 8. These findings can help guide clinicians in several ways. First, clinicians should consider re-evaluating treatment if a high severity patient has not shown meaningful PGD symptom improvement by week 8. Second, among patient characteristics, higher baseline depression severity and grief-related functional impairment were the most important clinical factors associated with decreased response to PGDT and/or citalopram. Previous applications of machine learning methods have routinely identified baseline depression severity as a top discriminative factor, as in the case of predicting responders to citalopram (Chekroud et al., 2016) and differential antidepressant efficacy (Chekroud et al., 2017; Chen et al., 2023) for MDD or in the development of precision treatment rules to optimize psychotherapy selection for PGD (Argyriou et al., 2024). Third, our analyses indicated, consistent with the primary study (Shear et al., 2016), that patients receiving PGDT were more likely to show symptom improvement.

Although limited in power, potential unmeasured factors, and the lack of an independent validation sample to confirm generalizability (Chekroud et al., 2024), these findings suggest individual and treatment factors that may impact treatment response for PGD. Findings should be viewed as useful associations in need of replication (Chekroud et al., 2024) rather than definitive causal relationships or specific mechanistic information. Instead, our results serve as accessible proxy information of response probability based on clinical profiles. Such response prediction data can support the development of decision support tools (e.g., prognostic risk calculators) that translate routine assessments within clinical practice into actionable tools to effectively estimate the likelihood of whether a specific patient will respond to a particular treatment.

## Conclusions

In this secondary analysis of an RCT of citalopram with and without PGDT, machine learning analyses identified depression severity and grief-related functional impairment as the most important baseline factors associated with differential response and replicated the effects of PGDT on outcomes from the primary study (Shear et al., 2016). Further, response trajectories significantly differed by 8 weeks, suggesting that therapeutic approaches may need to be adapted if reductions in grief severity are not observed by 8 weeks. We extend previous findings suggesting baseline factors may support personalizing and improving treatment outcomes for PGD. Future studies are needed to validate these results and their clinical applicability. Of particular interest will be the examination of individual PGD symptoms, moving beyond symptoms sum scores to account for their complex interactions (Maccallum et al., 2017; Malgaroli et al., 2018). This line of research would benefit from sophisticated data collection approaches (Galatzer-Levy and Onnela, 2023), which include the direct examination of loss narrative to directly assess symptoms using Natural Language Processing (Malgaroli et al., 2023), and the use of advanced computational methods to analyze their temporal dynamics (Brindle et al., 2024). The ultimate goal of this more sophisticated computational research is to refine our models of the transition boundaries between culturally appropriate and maladaptive chronic grief reactions, generating insights into treatment mechanisms to facilitate recovery (Lunansky et al., 2024).

## CRediT authorship contribution statement

**Adam Calderon**: Conceptualization, Data curation, Formal analysis, Investigation, Methodology, Software, Validation, Visualization, Writing – original draft, Writing – review and editing. **Matthew Irwin**: Project administration, Writing – original draft, Writing – review and editing. **Naomi M. Simon**: Conceptualization, Data curation, Funding acquisition, Investigation, Methodology, Project administration, Resources, Supervision, Validation, Writing – original draft, Writing – review and editing. **M. Katherine Shear**: Conceptualization, Data curation, Funding acquisition, Investigation, Data curation, Resources, Supervision, Validation, Writing – review and editing, Project administration. **Christine Mauro**: Investigation, Data curation, Project administration, Writing – review and editing. **Sidney Zisook**: Conceptualization, Data curation, Funding acquisition, Investigation, Project administration, Resources, Supervision, Validation, Writing – review and editing. **Charles F. Reynolds III**: Conceptualization, Data curation, Funding acquisition, Investigation, Project administration, Resources, Supervision, Validation, Writing – review and editing. **Matteo Malgaroli**: Conceptualization, Data curation, Formal analysis, Investigation, Methodology, Project administration, Software, Supervision, Validation, Visualization, Writing – original draft, Writing – review and editing.

## Financial support

The study was supported by grants R01MH60783, R01MH085297, R01MH085288, R01MH085308, and P30 MH90333 from the National Institutes of Health and by grant LSRG-S-172-12 from the American Foundation for Suicide Prevention. MM research was supported by K23MH134068 from the National Institutes of Health. The content is solely the responsibility of the authors and does not necessarily represent the official views of the NIH.

## Role of the Funder/Sponsor

The funders (the National Institute of Mental Health and the American Foundation for Suicide Prevention) had no role in the design and conduct of the study; the collection, management, analysis, and interpretation of the data; the preparation, review, or approval of the manuscript; or the decision to submit the manuscript for publication.

## Conflict of Interest Disclosures

Dr. Shear reported grant funding from the US Department of Defense, Congressionally Directed Medical Research Programs and a contract from Guilford Press to write a book on grief. In the past 3 years Dr. Simon reports receiving grants from the National Institutes of Health (NIH), American Foundation for Suicide Prevention, Patient-Centered Outcomes Research Institute, Ananda Scientific and support from Cohen Veterans Network and MindMed; receiving personal fees from Genomind, Cerevel, Engrail, Praxis; receiving fees or royalties from Wiley (Deputy Editor Depression and Anxiety), Wolters Kluwer (UpToDate) and APA Publishing (Textbook of Anxiety, Trauma and OCD Related Disorders 2020); and having spousal stock from G1 Therapeutics and Zentalis outside the submitted work.

Dr. Reynolds reported receiving pharmaceutical support for National Institutes of Health– sponsored research studies from Bristol-Myers Squibb, Forest, Pfizer, and Lilly; receiving grants from the National Institute of Mental Health, National Institute on Aging, National Center for Minority Health Disparities, National Heart, Lung, and Blood Institute, Center for Medicare and Medicaid Services, Patient Centered Outcomes Research Institute, the Commonwealth of Pennsylvania, the John A. Hartford Foundation, National Palliative Care Research Center, Clinical and Translational Science Institute, and the American Foundation for Suicide Prevention; and serving on the American Association for Geriatric Psychiatry editorial review board. He has received an honorarium as a speaker from MedScape/WebMD. He is the coinventor (licensed intellectual property) of psychometric analysis of the Pittsburgh Sleep Quality Index PRO10050447, supported by the National Institutes of Health through grants P60 MD000207, P30 MH090333, UL1RR024153, and UL1TR000005 and the University of Pittsburgh Medical Center endowment in geriatric psychiatry.

## Data Availability

All data produced in the present study are available upon reasonable request to the authors

## Notes

### Clinical Trial

NCT01179568

### Clinical Protocols

https://shorturl.at/Ng4HB

### Author Declarations

NEW YORK STATE PSYCHIATRIC INSTITUTE - COLUMBIA UNIVERSITY DEPARTMENT OF PSYCHIATRY INSTITUTIONAL REVIEW BOARD

