## Supplementary Materials for "Depression is Associated with Treatment Response Trajectories in Adults with Prolonged Grief Disorder: A Machine Learning Analysis"

### Supplementary Online Content

**Appendix A.** Extended methods of the secondary analysis

**Appendix B.** Rationale for deviations from trial protocol

**Supplementary Figure 1.** Discriminatory power of final nested cross-validated elastic net model for classification of remitting trajectory groups using patients' characteristics (n = 333)

**Supplementary Figure 2.** Discriminatory power of the training model (inner-fold) of nested cross-validated elastic net model (n = 333)

**Supplementary Figure 3.** AUC-PR curve of an elastic net model based on single 5-fold and 10 times repeated 5-fold nested cross-validation (n = 333)

**Supplementary Figure 4.** Confusion matrix of final nested cross-validated elastic net model (n = 333)

**Supplementary Figure 5.** Binomial deviance based on lambda tuning for final nested cross-validated elastic net model

**Supplementary Figure 6.** Binomial deviance based on alpha tuning for final nested cross-validated elastic net model

**Supplementary Figure 7.** Number of non-zero coefficients based on alpha tuning for final nested cross-validated elastic net model

**Supplementary Figure 8.** Conditional latent growth mixture model with known treatment assignment (n = 333)

**Supplementary Figure 9.** SHAP waterfall plot of final nested cross-validated elastic net model (n = 333)

**Supplementary Figure 10.** Discriminatory power of a nested cross-validated elastic net model based on only patients who received PGDT (n = 174)

**Supplementary Figure 11.** Variable importance and potential directionality based on SHAP for a nested cross-validated elastic net model based on only patients who only received PGDT (n = 174)

**Supplementary Figure 12.** Discriminatory power of a nested cross-validated elastic net model based on only patients who received CIT (n = 159)

**Supplementary Figure 13.** Variable importance and potential directionality based on SHAP for a nested cross-validated elastic net model based on only patients who only received CIT (n = 159)

**Supplementary Table 1.** Growth mixture model fit indices

**Supplementary Table 2.** Final coefficients and Shapley values of baseline variables retained in final elastic net model (n = 333)

**Supplementary Table 3.** Model performance metrics of final nested cross-validated elastic net model (n = 333)

This supplementary material has been provided by the authors to give readers additional information about their work.

**Appendix A.** Extended methods

*Latent Growth Mixture Modeling (LGMM)*

We explored intercept, slope, and quadratic parameters as either random or fixed effects. In the final models, intercept variances were allowed to be freely estimated, whereas linear slope variances were fixed, and quadratic parameters were nonsignificant and thus removed to facilitate model convergence. Model solutions from one to five classes were compared with the optimal number of classes determined by comparing a combination of indices for improvements of fit (Nylund et al., 2007): Akaike information criterion, Bayesian information criterion, sample-size-adjusted Bayesian information criterion, relative entropy, Lo-Mendell-Rubin-adjusted likelihood ratio test, and parametric bootstrapped likelihood ratio test. A solution is deemed to have the best relative fit based on lower information criteria values, significant likelihood ratio tests, and higher entropy (Jung and Wickrama, 2008). Solutions with small sample sizes within a class were excluded because these tend to be unstable and difficult to replicate (Nylund-Gibson and Choi, 2018). In addition, explanatory properties, theoretical coherence, and parsimony are also considered to determine the optimal number of classes (Muthén, 2003).

*Machine Learning model development and internal validation*

The outcome of the logistic regression with elastic net regularization (elastic net) model was a binary classification of longitudinal symptom trajectories of remitting grief symptoms versus non-remitting trajectories, as initially identified by LGMM. Candidate features constituted patient-level characteristics collected prior to the RCT. For the current study, sudden death variable was operationalized as death occurring due to illness in less than one month, accident, murder, suicide or other vs. other type of loss; the distinction was made to assess the suddenness of death experienced. Violent death was operationalized as death constituting accident, murder, suicide, or other vs. other type of loss; the distinction was made to assess the violent nature of death experienced. Suicidal ideation, current MDD, GAD, and PTSD diagnoses were operationalized as caseness vs. absence. Using one hot encoding (i.e., dummy codes), sleep disturbance, spousal loss, child loss, sudden death, and violent death were dichotomized to represent caseness vs. absence. Moreover, race was dichotomized to represent white vs. other, gender was dichotomized to represent male vs. female, ethnicity was dichotomized to represent Hispanic vs. non-Hispanic, and level of education was dichotomized to represent the presence vs. absence of a bachelor’s degree or higher.

To optimize the probability of unbiased results, we utilized a state-of-the-art method termed $k \times l$-*fold* nested cross-validation to prevent over-fitting (Lewis et al., 2023). Unlike traditional machine learning train/test splits, nested cross-validation utilizes the whole dataset by partitioning it into outer and inner folds. Put briefly, the inner-fold cross-validation is used to tune optimal hyperparameters for models (e.g., select optimal shrinkage parameters for the elastic net model). Then, the model is fitted on the whole outer training fold and tested on the inner fold using the left-out data from the outer fold. This is repeated across all outer folds, and the pooled unseen test predictions from the outer folds are compared against the true results for the outer test folds, and the predicted probabilities are concatenated. Thus, providing measures of accuracy across the whole dataset. Once the performance of the elastic net model is evaluated across all test partitions, a final step of model development is performed by retraining an elastic net model with the optimal set of hyperparameters and selected predictors using the whole dataset, which can be used for prediction with external data. Notably, nested cross-validation has been shown to reduce cross-validation bias and replicate error estimates parallel to those obtained from independent external validation (Krstajic et al., 2014; Varma and Simon, 2006). For the current study, we utilized 5-fold nested cross-validation for the outer and inner folds.

Missing data was handled using random forest imputation via the missRanger R package (Mayer, 2017). Random forest imputation stands as a robust non-parametric approach in handling mixed data types containing continuous and categorical variables and capturing potential complex interactions and non-linear relation structures (Pantanowitz and Marwala, 2009; Stekhoven and Bühlmann, 2012). Moreover, given that the principal objective of machine learning is to optimize the performance of models, we also addressed class imbalance by applying the synthetic minority oversampling technique (Chawla et al., 2002), which undersamples the high-frequency class and oversamples the low-frequency class to generate a more balanced structure for the outcome. Class imbalance occurs when machine learning models favor predictions from the majority class and ignore the minority class, given preferences for high accuracy, even if purely by chance (Jacobucci and Li, 2022).

*Model performance evaluation*

For interpretive purposes, we adopted the following framework: AUC = .50 reflects no discrimination, .70 ≤ AUC ≤ .80 reflects acceptable discrimination, and AUC ≥ .80 reflects excellent discrimination. We also evaluated standard ROC metrics using a threshold of 0.50. Namely, accuracy (i.e., number of corrected classified cases with respect to all cases), balanced accuracy (i.e., accuracy that ensures both minority and majority classes are equally important ), recall (i.e., sensitivity; the proportion of well predicted cases with respect to the total number of observed cases for the positive class), specificity (i.e., selectivity; represents the proportion of well classified negative values with respect to the total number of actual negatives), precision (i.e., positive predictive value; the proportion of well classified cases with respect to the total of cases predicted with the true class), F1 score (harmonic mean of precision and recall), negative predictive value (i.e., proportion of true negatives among all predicted negatives), prevalence (i.e., proportion of the total cases that are positive), false discovery rate (proportion of false positives among all predicted positives), no information rate (i.e., proportion of observations that fall into the majority class), geometric mean (balanced performance of both majority and minority classes), Fowlkes-Mallows Index (i.e., the similarity between predicted and observed), Cohen’s kappa coefficient (i.e., inter-rater reliability normalized by the possibility of agreement by chance), Youden’s J-Index (i.e., the maximum vertical distance between ROC curve and diagonal line that maximizes the difference between true positive rate and false positive rate), Jaccard’s Index (i.e., the size of the intersection divided by the size of the union between two label sets), Markedness (i.e., the trustworthiness of positive and negative predictions by model), Positive likelihood ratio (i.e., the odds of obtaining a positive prediction for actual positives), Negative likelihood ratio (i.e., odds of obtaining a negative prediction for actual positives relative to the probability of actual negatives of obtaining a negative prediction), and diagnostic odd ratio (i.e., the odds of a positive case obtaining a positive prediction result with respect to the odds of actual negatives obtaining a positive result).

**Appendix B.** Rationale for deviations from trial protocol

The current secondary analysis deviated from the main outcome paper with its use of ICG scores to create trajectories that better differentiate subgroups of patients as opposed to the Clinical Global Impression scale (CGI).(Guy, 1976) We based this decision on technical (e.g., larger numerical range) and clinical elements (e.g., ICG is more directly aligned with PGD symptoms), given the current study's interest in utilizing grief severity to determine by which week individuals would be considered non-responders (week 8).

**Figure S1**

*Discriminatory power of final nested cross-validated elastic net model for classification of remitting trajectory groups using patients' characteristics (n = 333)*

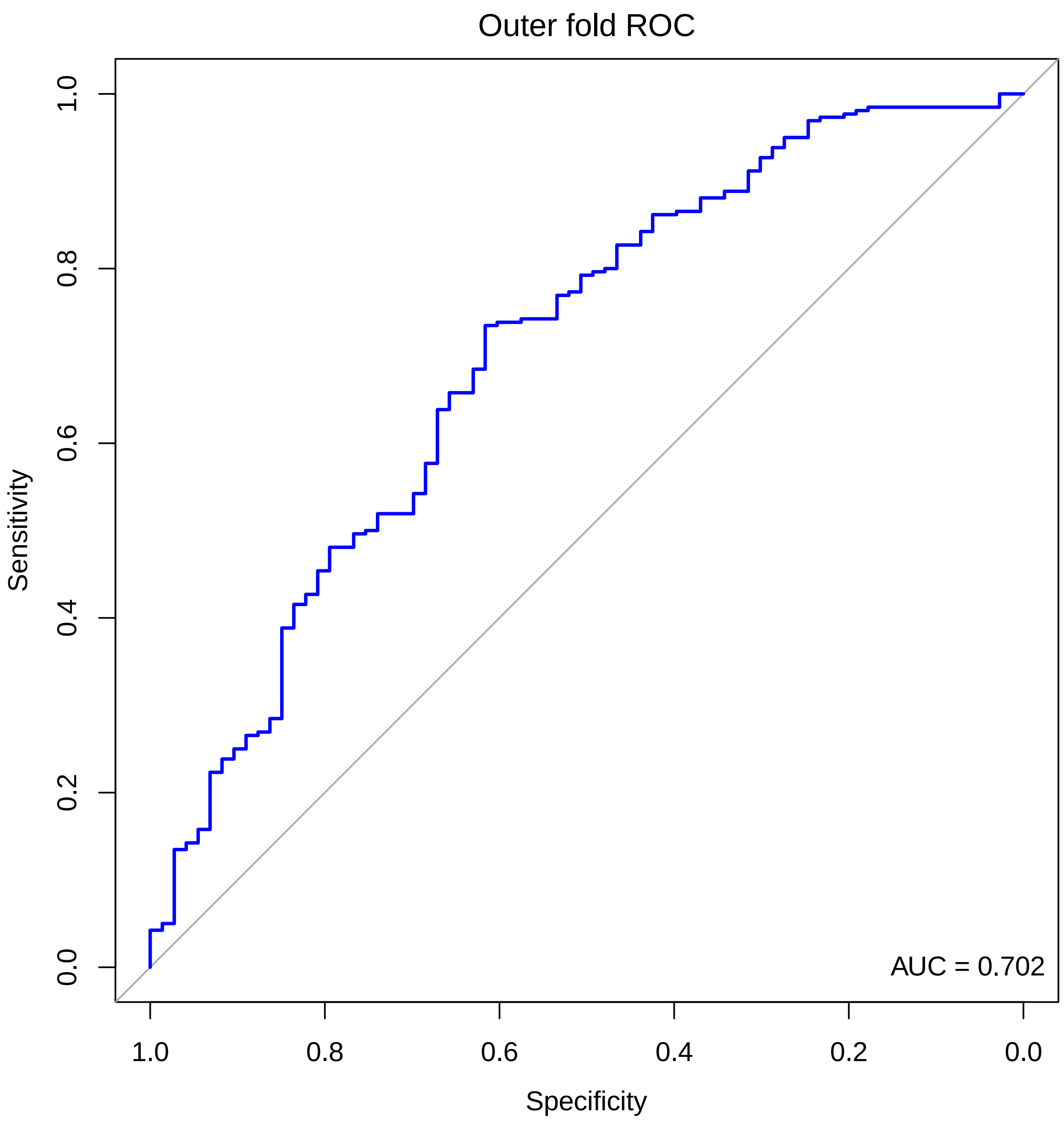


*Note*. Abbreviations: ROC, receiver operating characteristic; AUC, area under the ROC curve. 5-fold nested cross-validation was applied for outer and inner folds.

**Figure S2**

*Discriminatory power of the training model (inner-fold) of nested cross-validated elastic net model (n = 333)*

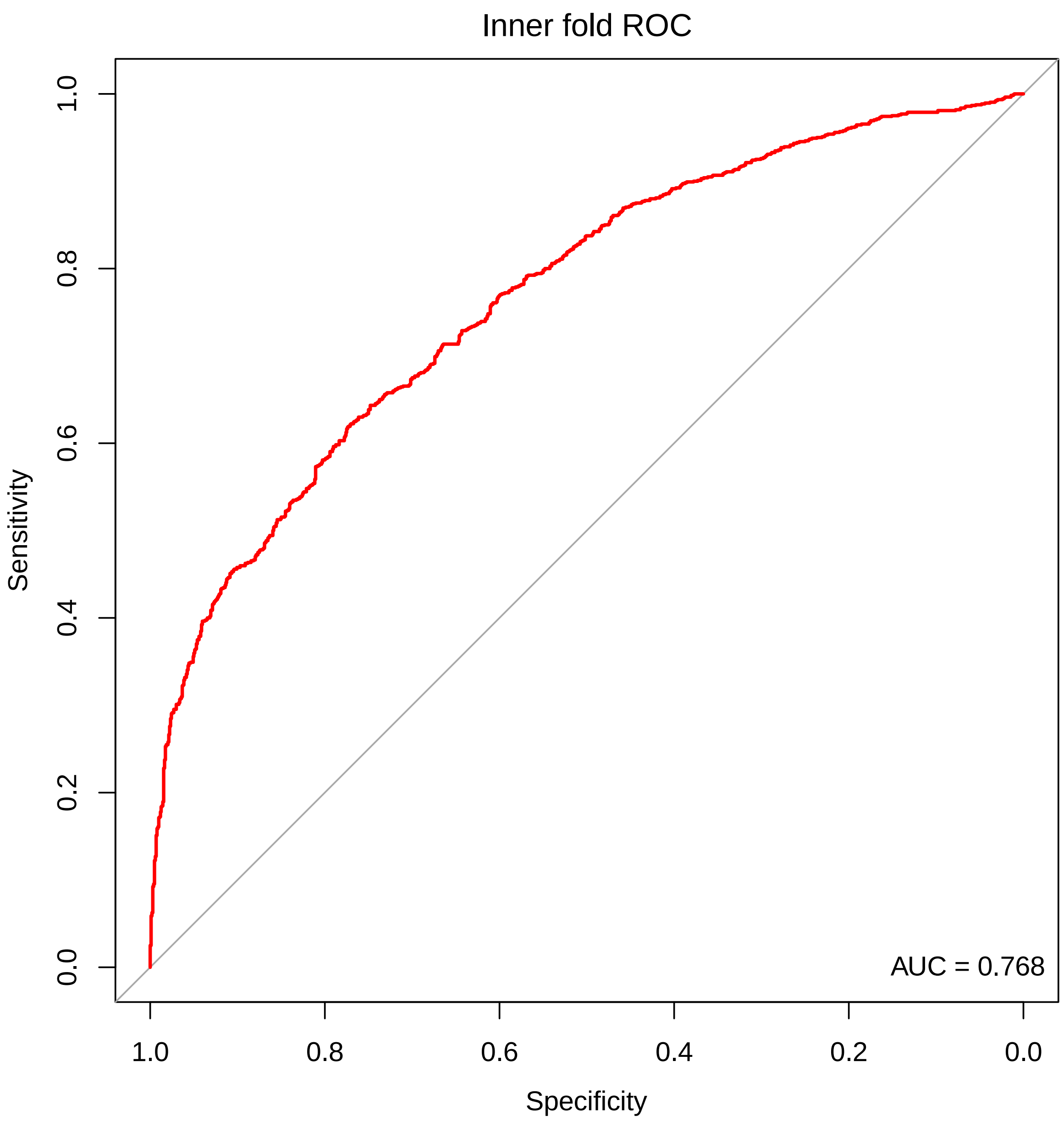


*Note*. Abbreviations: ROC, receiver operating characteristic. Supplementary Figure S2 reports the inner fold cross-validation which was used to tune optimal hyperparameters for the elastic net model. Accordingly, it does not reflect final performance as it has not been subsequently tested on the left-out data from the outer fold. Therefore, Supplementary Figure S2 solely represents the discriminatory power of the "training model." During model training, 5-fold nested cross-validation was applied.

**Figure S3**

*AUC-PR curve of an elastic net model based on single 5-fold and 10 times repeated 5-fold nested cross-validation (n = 333)*

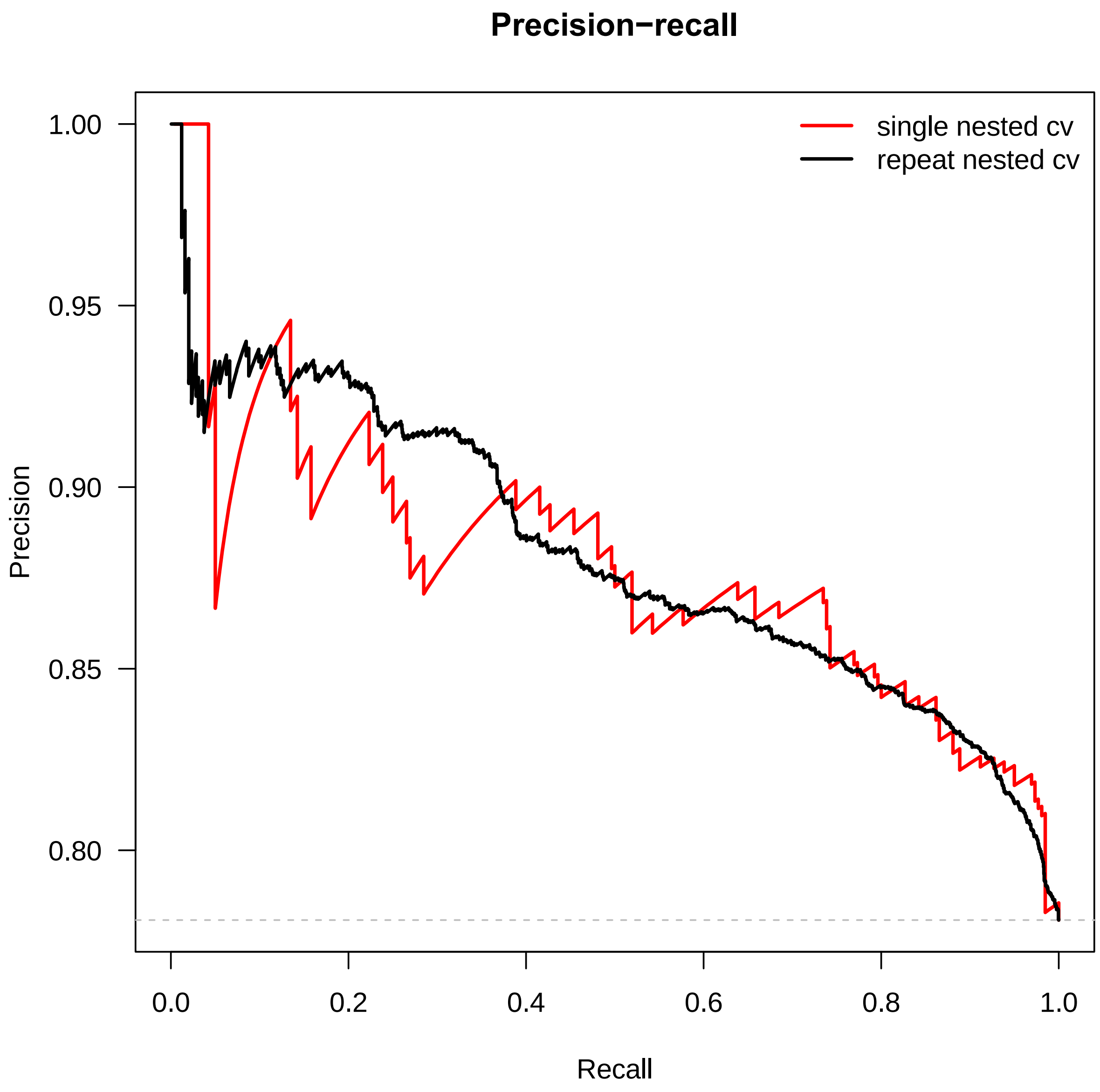


*Note*. Abbreviations: AUC-PR, area under the precision recall curve; single nested cv, 5-fold nested-cross validation performed once; repeat nested cv, 5-fold nested-cross validation repeated 10 times. For 10 times repeated 5-fold nested cross-validation, test fold predictions are concatenated across all repeats. Supplementary Figure S3 illustrates a smoother AUC-PR compared to the AUC-PR curve from a single model. Overall, accurate overall estimates of performance may be seen for both nested cross-validation approaches.

**Figure S4**

*Confusion matrix of final nested cross-validated elastic net model (n = 333)*

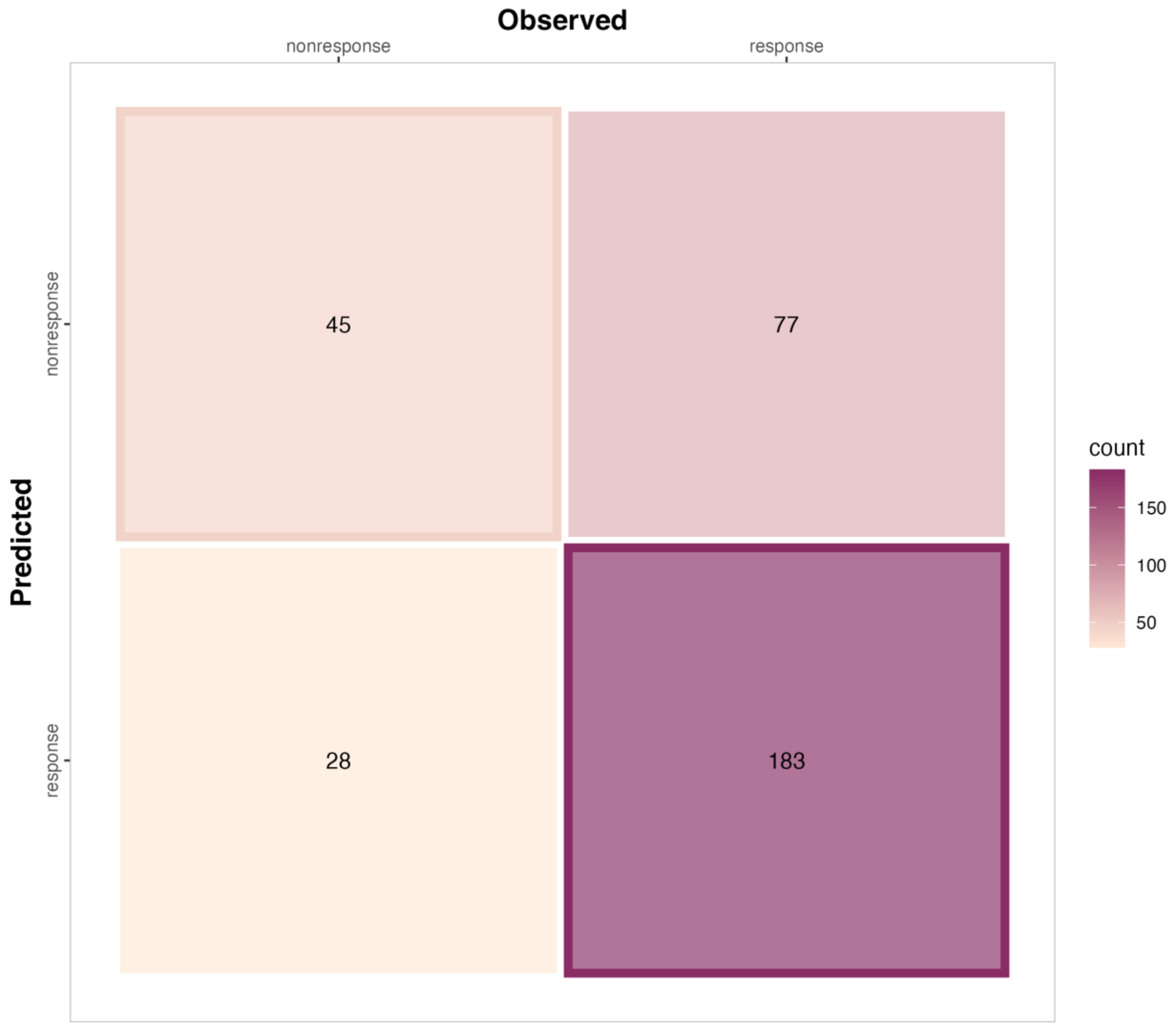


*Note.* Abbreviations: response, longitudinal symptom trajectories of remitting grief symptoms; nonresponse, longitudinal symptom trajectories of non-remitting grief symptoms.

**Figure S5**

*Binomial deviance based on lambda tuning for final nested cross-validated elastic net model*

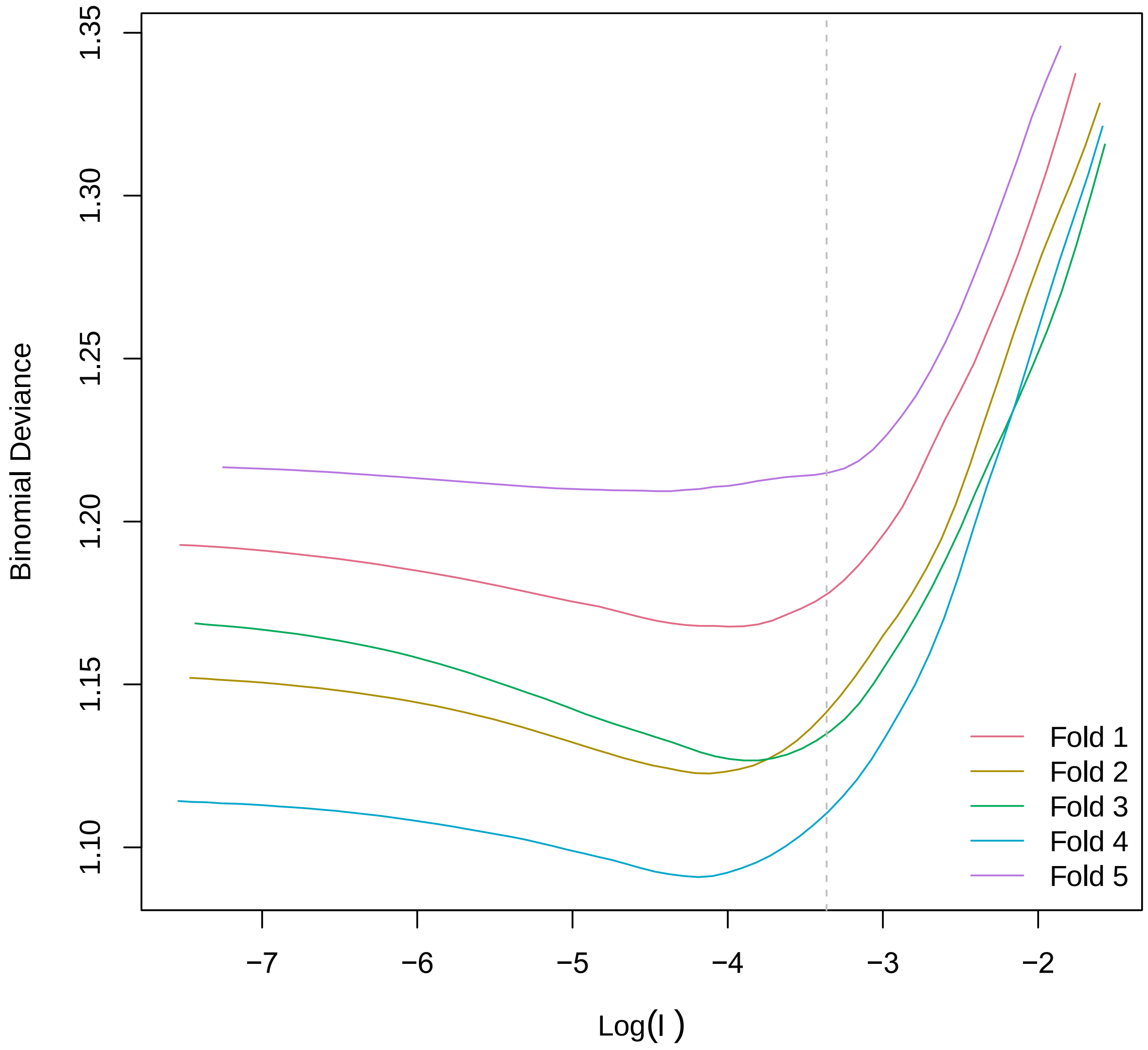


Note. Abbreviations: λ, lambda.

**Figure S6**

*Binomial deviance based on alpha tuning for final nested cross-validated elastic net model*

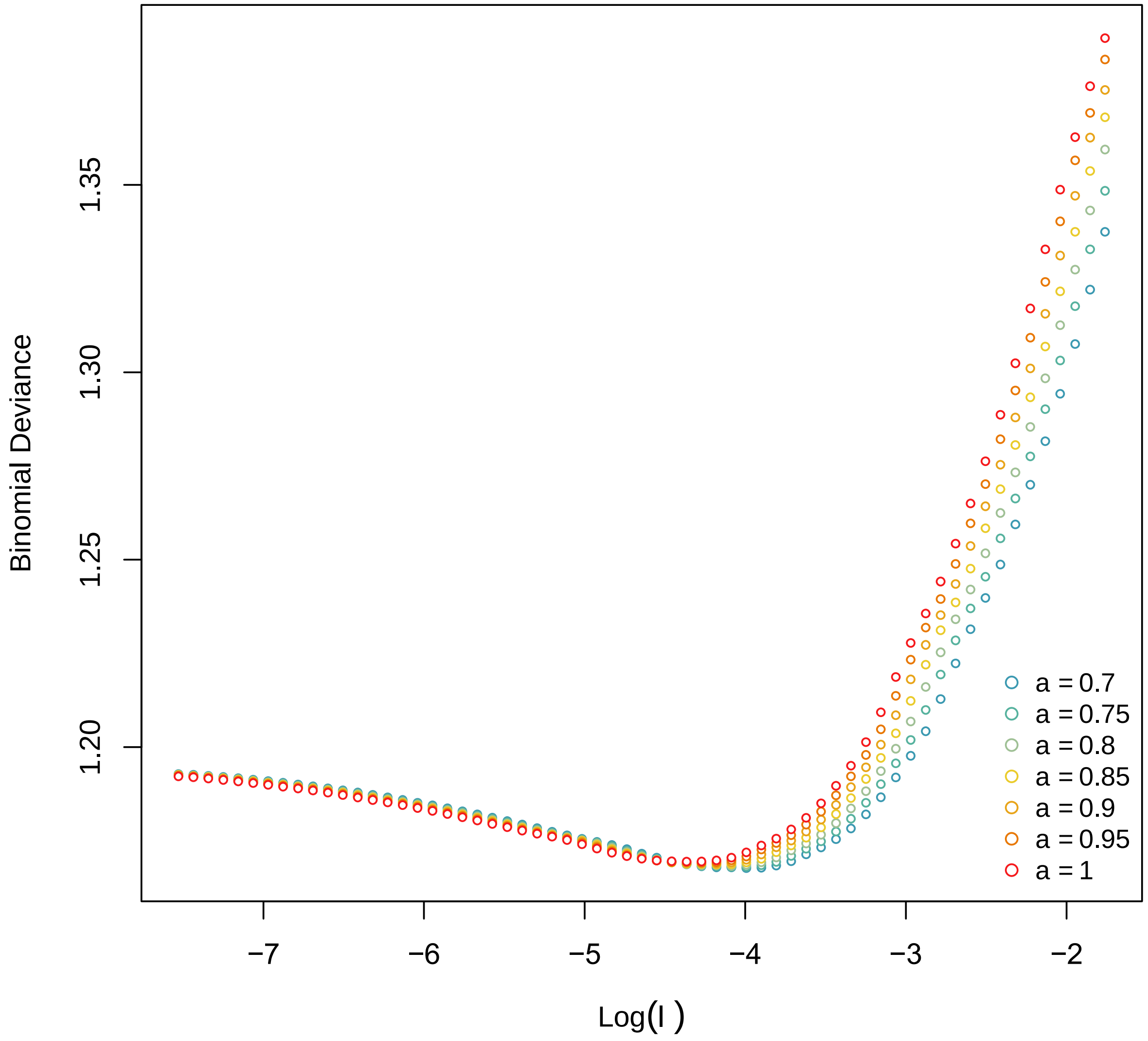


Note. Abbreviations: α , alpha.

**Figure S7**

*Number of non-zero coefficients based on alpha tuning for final nested cross-validated elastic net model*

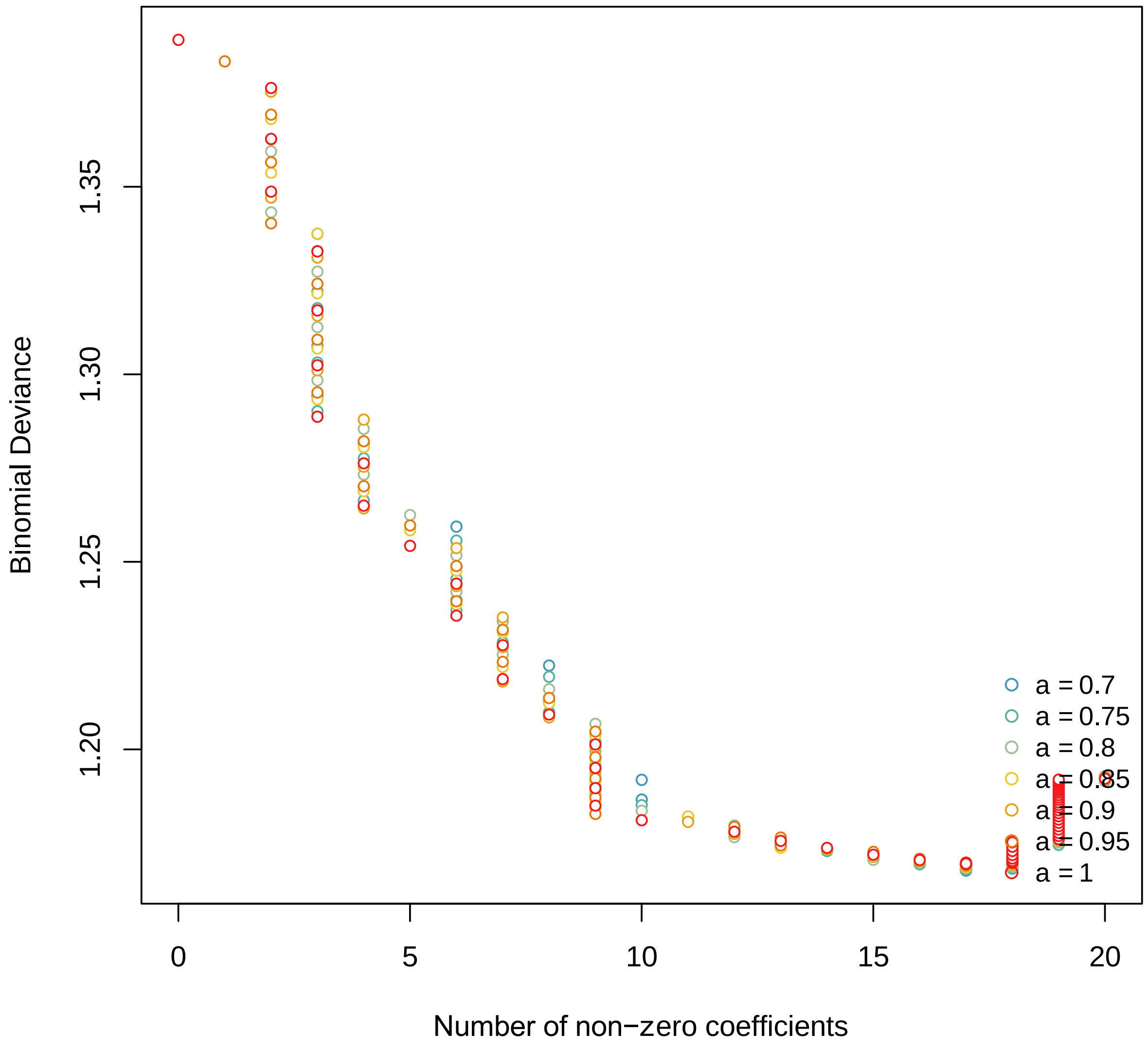


Note. Abbreviations: α, alpha.

**Figure S8**

*Conditional latent growth mixture model with known treatment assignment (n = 333)*

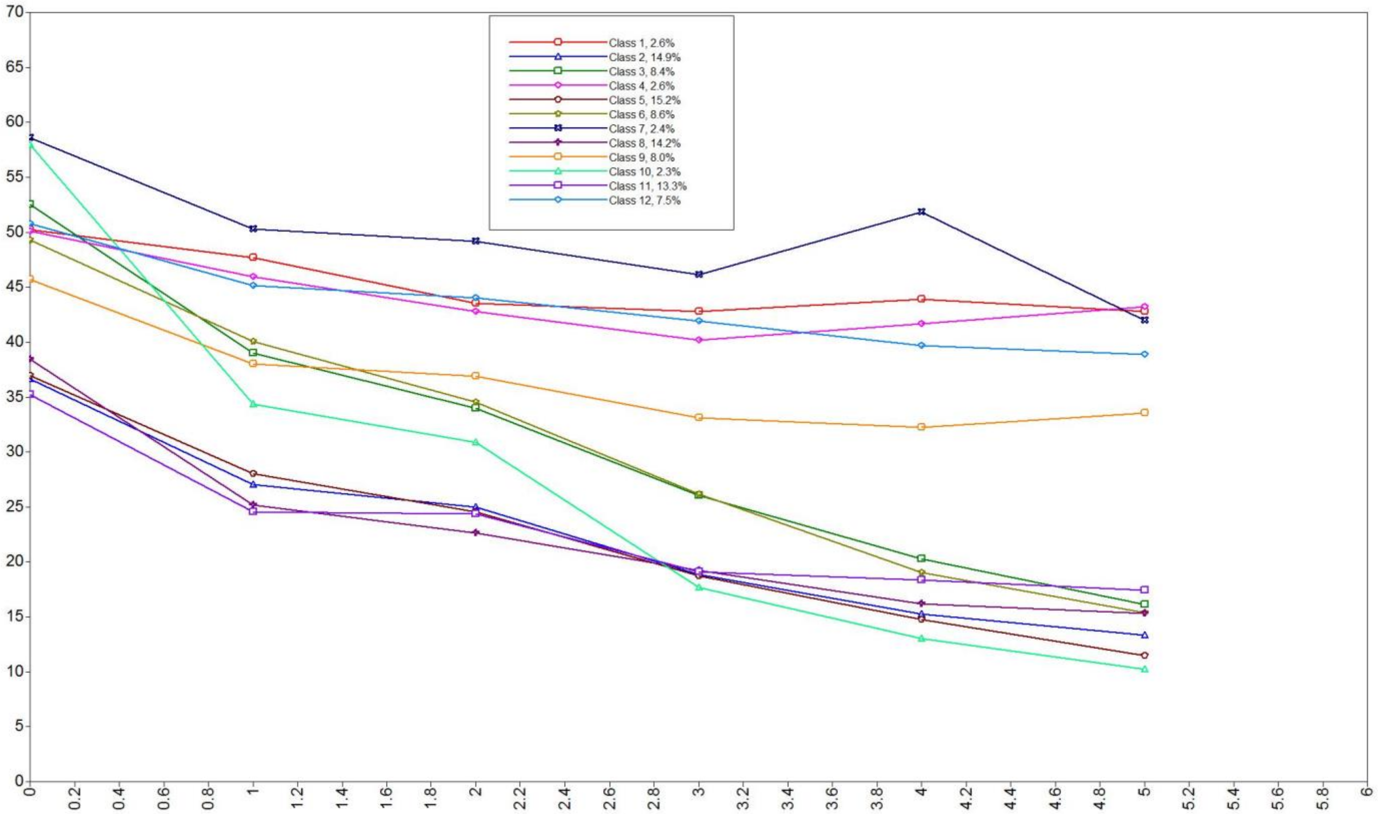


*Note.* Abbreviations: CIT, citalopram; PGDT, Prolonged Grief Disorder therapy; PLA, pill placebo. As a moderation analysis, we ran a supplemental trajectory analysis with treatment assignment assigned as a known class. Supplementary Figure S8 distinguishes differences in proportions within trajectory class based on assigned treatment groups. Class 1 (2.6%): CIT + PGDT, lesser severity responders. Class 2 (14.9%): CIT + PGDT, greater severity responders. Class 3 (8.4%): CIT + PGDT, non-responders. Class 4 (2.6%): PLA + PGDT, lesser severity responders. Class 5 (15.2%): PLA + PGDT, greater severity responders. Class 6 (8.6%): PLA + PGDT, non-responders. Class 7 (2.4%): CIT + No PGDT, lesser severity responders. Class 8 (14.2%): CIT + No PGDT, greater severity responders. Class 9 (9.0%): CIT + No PGDT, non-responders. Class 10 (2.3%): PLA + No PGDT, lesser severity responders. Class 11 (13.3%): PLA + No PGDT, greater severity responders. Class 12 (7.5%): PLA + No PGDT, non-responders. Overall, results demonstrated no significant differences in the proportions between the trajectory class and treatment groups.

**Figure S9**

*SHAP waterfall plot of final nested cross-validated elastic net model (n = 333)*

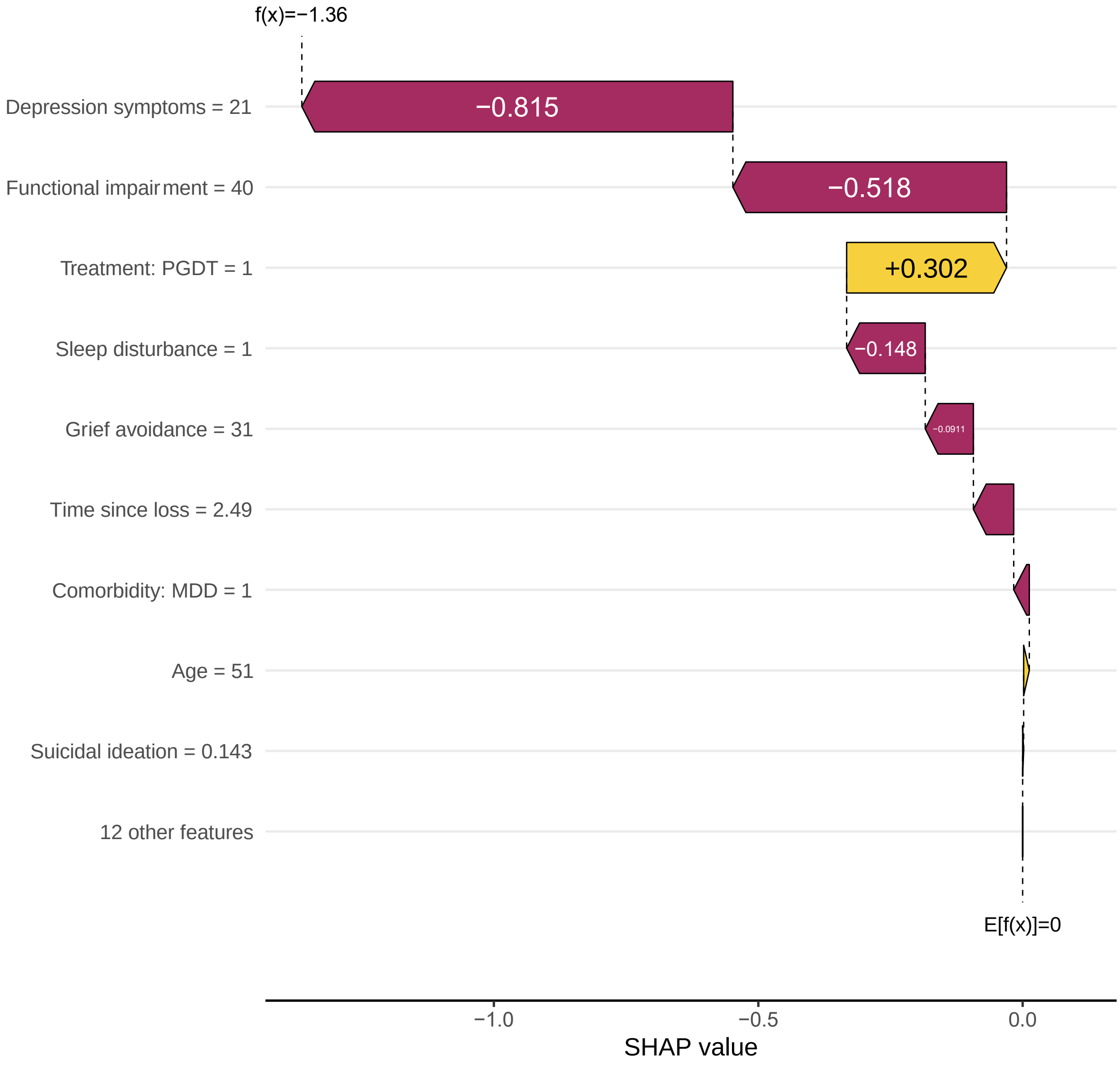


*Note.* Abbreviations: SHAP, SHapley Additive explanation. SHAP waterfall plots illustrate how candidate features contributed to the model's output. Features are sorted by the magnitude of their SHAP values with the smallest magnitude features grouped together.

**Figure S10**

*Discriminatory power of a nested cross-validated elastic net model based on only patients who received PGDT (n = 174)*

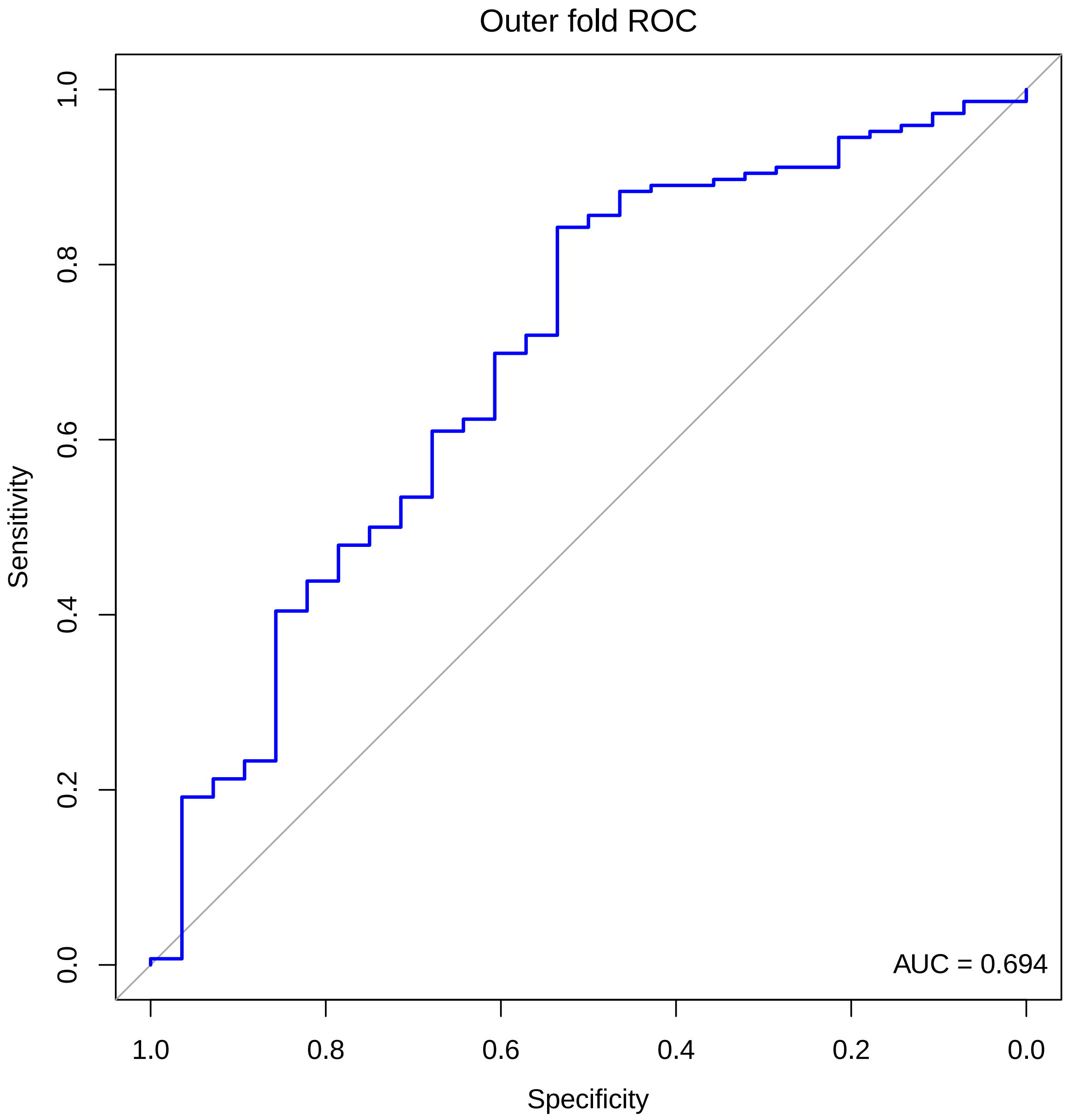


*Note*. Abbreviations: PGDT, Prolonged Grief Disorder therapy; ROC, receiver operating characteristic; AUC = area under the ROC curve. Supplementary Figure S10 represents a supplemental elastic net model utilizing parallel methods to the final nested cross-validated elastic net model but was trained only on the sub-sample of patients who only received PGDT. Supplementary Figure S10 depicts discriminatory power for the classification of remitting trajectory groups using patients' characteristics. 5-fold nested cross-validation was applied for outer and inner folds.

**Figure S11**

*Variable importance and potential directionality based on SHAP for a nested cross-validated elastic net model based on only patients who only received PGDT (n = 174)*

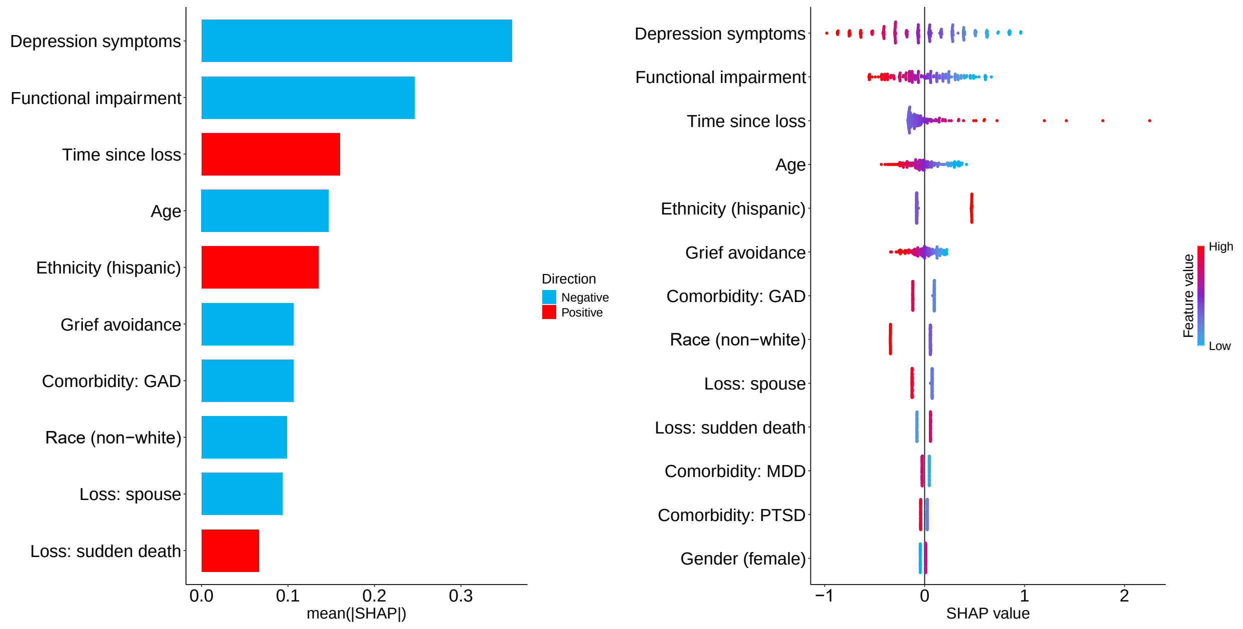


*Note.* Abbreviations: PGDT, Prolonged Grief Disorder therapy; SHAP, SHapley Additive exPlanations. Supplementary Figure S11 represents feature importance derived from a supplemental elastic net model that was trained only on a sub-sample of patients who only received PGDT. The bar plot (left) shows the mean absolute SHAP value per feature. Features are first sorted by their global impact (*y* axis) and are arranged in descending order based on the magnitude of their total contribution. The larger the SHAP value, the stronger the contribution within the model in discriminating between response trajectories. The (x-axis) represents the probability for treatment response as log odds. The bee swarm plot (right) further illustrates the potential directionality of the associations between features and classification by representing every individual in the sample as a dot denoting the attribution value for each feature from low (blue) to high (red).

**Figure S12**

*Discriminatory power of a nested cross-validated elastic net model based on only patients who received CIT (n = 159)*

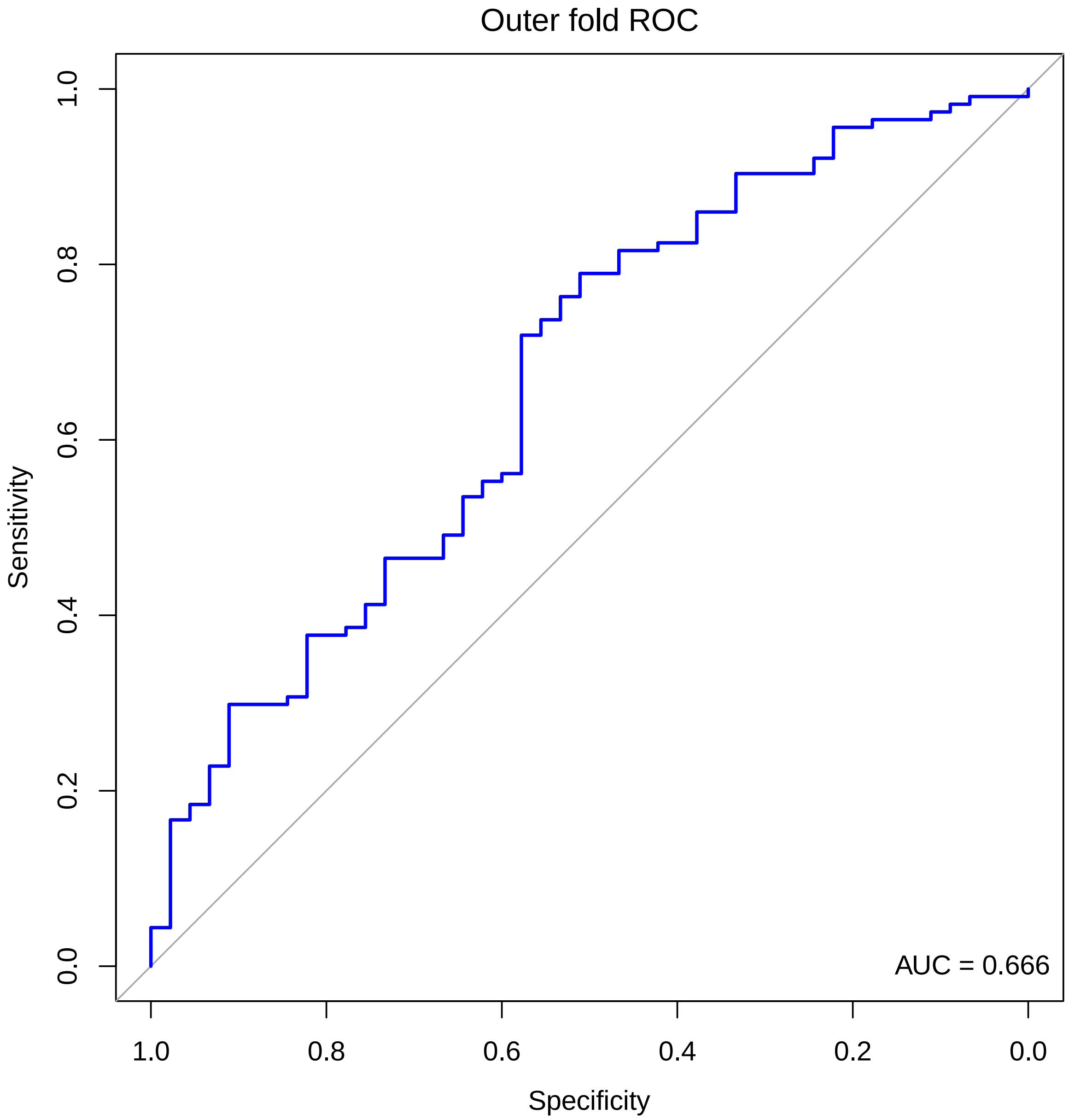


*Note*. Abbreviations: CIT, citalopram; ROC, receiver operating characteristic; AUC, area under the ROC curve. Supplementary Figure S12 represents a supplemental elastic net model utilizing parallel methods to the final nested cross-validated elastic net model but was trained only on the sub-sample of patients who only received CIT. Supplementary Figure S12 depicts discriminatory power for the classification of remitting trajectory groups using patients' characteristics. 5-fold nested cross-validation was applied for outer and inner folds.

**Figure S13**

*Variable importance and potential directionality based on SHAP for a nested cross-validated elastic net model based on only patients who only received CIT (n = 159)*

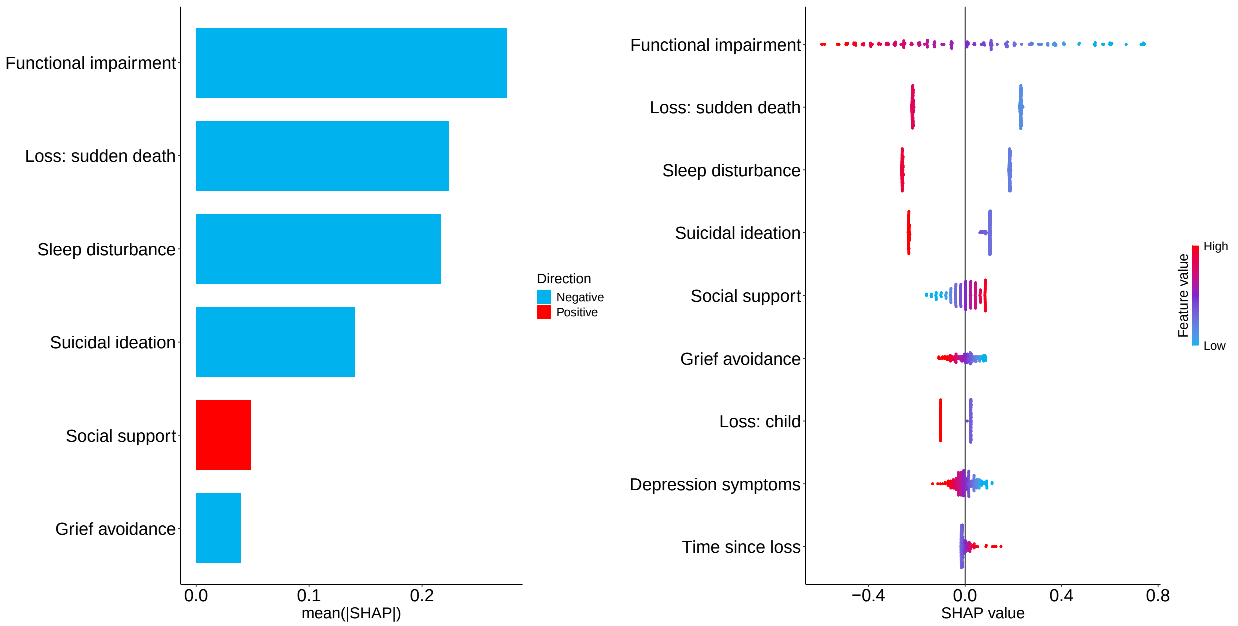


*Note.* Abbreviations: CIT, citalopram; SHAP, SHapley Additive exPlanations. Supplementary Figure S13 represents feature importance derived from a supplemental elastic net model that was trained only on a sub-sample of patients who only received CIT. The bar plot (left) shows the mean absolute SHAP value per feature. Features are first sorted by their global impact (*y* axis) and are arranged in descending order based on the magnitude of their total contribution. The larger the SHAP value, the stronger the contribution within the model in discriminating between response trajectories. The (x-axis) represents the probability for treatment response as log odds. The bee swarm plot (right) further illustrates the potential directionality of the associations between features and classification by representing every individual in the sample as a dot denoting the attribution value for each feature from low (blue) to high (red).

**Table S1**

*Growth mixture model fit indices*

| Statistics | 1 Class | 2 Classes | 3 Classes | 4 Classes | 5 Classes |
|---|---|---|---|---|---|
| Akaike Information Criteria | 12594.904 | 12375.317 | 12335.881 | 12306.959 | 12294.372 |
| Bayesian Information Criteria | 12640.602 | 12436.247 | 12412.044 | 12398.354 | 12401.000 |
| Sample-Size Adjusted BIC | 12602.537 | 12385.494 | 12348.603 | 12322.225 | 12312.182 |
| Entropy | - | 0.774 | 0.796 | 0.779 | 0.815 |
| Lo-Mendell-Rubin Adjusted LRT | - | 218.196 | 45.478 | 35.399 | 19.737 |
| *P-value* | - | *<0.05 or = 0.0000* | *<0.05 or = 0.0158* | *<0.05 or = 0.0177* | *<0.05 or = 0.0027* |
| Smallest sample size (n) | 333 | 130 | 60 | 42 | 2 |

*Note*. Model fit information for nested unconditional latent growth mixture models with increasing class size from 1 to 5 trajectories are provided. Information indices and likelihood tests showed improved fit as the number of classes increased from one to five solutions; however, entropy decreased with a four-class solution. Moreover, the addition of a fifth class resulted in a class constituting two individuals, making the model less parsimonious and interpretable. Consequently, the best fitting model was a three-class solution which had the lowest information criteria (Akaike information criterion = 12335.881, Bayesian information criterion = 12412.044, Sample-size adjusted Bayesian information criterion = 12348.603, Lo-Mendell-Rubin adjusted likelihood ratio test) and the highest relative entropy (0.796) and *p*-value (.0158).

**Table S2**

*Final coefficients and Shapley values of baseline variables retained in final elastic net model (n = 333)*

| Variable | *B* | SHAP |
|---|---|---|
| **Intercept** | 2.479 | - |
| **Treatment: PGDT** | 0.636 | 0.317 |
| **Sleep disturbance** | -0.274 | 0.135 |
| **Depression symptoms** | -0.107 | 0.346 |
| **Comorbidity: MDD** | -0.088 | 0.039 |
| **Time since loss** | 0.030 | 0.136 |
| **Functional impairment** | -0.028 | 0.236 |
| **Suicidal ideation** | -0.012 | 0.005 |
| **Grief-related avoidance** | -0.010 | 0.106 |
| **Age** | -0.003 | 0.044 |

*Note.* Abbreviations: *B*, final coefficient; SHAP, SHapley Additive exPlanations. SHAP values represent the average of all the marginal contributions to all possible combinations. Final nested cross-validated elastic net model employed an alpha of 1.000 and a lambda of .034.

**Table S3**

*Model performance metrics of final nested cross-validated elastic net model (n = 333)*

| Metric | *M* |
|---|---|
| **AUC** | .702 |
| **AUC via trapezoid approach** | .767 |
| **Area under precision-recall curve** | .874 |
| **Accuracy** | .684 |
| **Balanced accuracy** | .660 |
| **Specificity** | .616 |
| **Precision (positive predictive value)** | .867 |
| **Negative predictive value** | .383 |
| **Recall (sensitivity)** | .703 |
| **F1 score** | .777 |
| **Adjusted F-score** | .778 |
| **Geometric mean** | .658 |
| **Cohen's kappa coefficient** | .258 |
| **Matthews correlation coefficient (phi-coefficient)** | .275 |
| **Fowlkes-Mallows index** | .781 |
| **Bookmaker informedness (Youden's index)** | .320 |
| **Critical success index (Jaccard's index)** | .714 |
| **Markedness** | .236 |
| **Positive likelihood ratio** | 1.83 |
| **Negative likelihood ratio** | .48 |
| **Diagnostic odd ratio** | 3.81 |
| **False positive rate** | .368 |
| **False negative rate** | .296 |
| **False detection rate** | .132 |
| **False omission rate** | .631 |
| **Prevalence** | .780 |
| **Prevalence threshold** | .483 |
| **No information rate** | .780 |
| **Events per variable** | 12.38 |
| **Expected calibration error** | .297 |
| **Maximum calibration error** | .072 |
| **Integrated calibration index** | .315 |

Note. *M* = Arithmetic mean. AUC = Area under the ROC Curve.
